# Assessment and Utilization of Patient-Reported Outcomes Measures to Improve Cardiovascular Disease Prevention

**DOI:** 10.1101/2019.12.11.19014399

**Authors:** Phoebe A Finneran, Tinamarie Sanborn, Xiao Guo, Kate C Traynor, Michael R. Jaff, Pradeep Natarajan

## Abstract

**Background:** The American Heart Association’s Life’s Simple 7 (LS7) consist of clinical factors and health-related behaviors associated with cardiovascular health. The prevalence of these health factors among patients seeking specialized cardiovascular care is unknown. We sought to quantify patient-reported cardiovascular risk among those seeking specialized cardiovascular care and implement tailored quality improvement interventions.

**Methods and Results:** Patients cared for by cardiovascular specialists at an academic medical center were surveyed with a modified LS7. We compared the prevalence of optimal health factors by atherosclerotic cardiovascular disease (ASCVD) status. Recent smokers or patients with ASCVD reporting non-adherence to antiplatelets/statins were contacted.

Surveys for 5,950 patients were collected during 2014 to 2016. The mean (SD) age was 64 (15) years, 2613 (44%) were women, and 3478 (58%) had ASCVD. Only 34 (0.6 %) achieved ideal status for all 7 factors, which did not differ by ASCVD status (*P*=0.48). Of 404 (6.8%) reporting recent smoking, 94/404 (23.3%) were successfully contacted, and 71/404 (17.6%) accepted support. Among those with ASCVD, 84 (2.4%) reported not taking a statin or antiplatelet without contraindications.

**Conclusions:** The prevalence of optimal health factors, including health-related behaviors, among patients cared for by cardiovascular specialists remains low. Assessment of patient-reported outcomes facilitates scalable interventions to improve cardiovascular disease prevention.

## Introduction

Cardiovascular diseases (CVD) remain the leading cause of death in the US and worldwide^1-3^. In the US, healthcare spending accounts for approximately 17% of the US economy, and CVD accounts for the largest single fraction of these expenditures^4^. The American Heart Association (AHA) defined 7 health-related behaviors and quantitative clinical variables deemed Life’s Simple 7 (LS7), related to smoking, weight, physical activity, diet, cholesterol levels, blood pressure, and fasting glucose that contribute to ideal cardiovascular health^5^. In 2010, the AHA outlined the promotion of cardiovascular health, through optimization of the LS7, as a key 2020 Impact Goal to reduce cardiovascular and stroke death^5^. These health factors are associated with greater life expectancy, decreased incidence of CVD, increased health-related quality-of-life, and lower individual healthcare costs^5^. However, the prevalence of ideal health factors generally among American adults is extremely low, with <1% achieving ideal status across all 7 health factors^6-11^. In a setting where patients are seeking care with cardiovascular and cerebrovascular specialists, the prevalence of these factors are unknown.

Clinical screening for metabolic risk factors and promoting strategies for behavior modification in concert with pharmacotherapy where appropriate, are key strategies for cardiovascular disease prevention^12,13^. Cardiovascular disease specialists are uniquely positioned to systematically assess modifiable risk factors via patient-reported outcomes and intervene to improve clinical outcomes^14^. In epidemiologic cohorts, a high prevalence of ideal health-related behaviors and optimally controlled risk factors are associated with reduced future cardiovascular disease risk but <3% are optimal for all factors^15-17^. The prevalence of self-reported cardiovascular health-related behaviors, are poorly defined in the health-care setting, let alone outpatient practices addressing cardiovascular diseases.

We conducted a quality improvement initiative within a single large academic medical center to determine the prevalence of optimal and suboptimal health factors amongst patients seeking care from cardiovascular disease specialists, and developed interventions to address suboptimal health-related behaviors. We observed that, even among patients cared for by cardiovascular disease specialists, optimal status across ideal health factors and behaviors is very low. We also demonstrate a framework for using patient-reported outcomes to address high risk cardiovascular health-related behaviors.

## Methods

We consecutively approached patients cared for by cardiovascular disease specialists at the Massachusetts General Hospital (MGH), a large academic medical center, in outpatient clinical settings. Outpatient clinics included the women’s heart health program, preventive cardiology, interventional cardiology, vascular surgery, vascular medicine, and stroke neurology. Patients were surveyed between May 2014 and March 2016. All patients seeking care were approached and those agreeable to fill out the questionnaire in English participated. This project was a quality improvement initiative and thereby exempt from oversight by the Partners Healthcare Institutional Review Board.

An electronic tablet-based survey (“My Health Check”) was developed using Tonic’s Patient Reported Outcomes platform (https://tonicforhealth.com/patientreportedoutcomes). In addition to the American Heart Association’s LS7 (S1 Table), we assessed for the presence of cardiovascular diseases and adherence to key medications (S1 Text). Patients were provided an electronic tablet to fill out the survey while waiting for physicians in waiting rooms or exam rooms.

To maximize accuracy of self-report of cholesterol, glucose, and blood pressure values, patients were provided a paper copy of their most recent lab values and blood pressure measurements; patients reconciled this report with values potentially ascertained in other settings (e.g. laboratory tests performed at another facility, home blood pressure measurements, etc). A personalized report was displayed on the iPad, printed for the patient, and uploaded into the electronic health record (S1 Fig, S1 Text). Ideal, intermediate, or poor status for each of the ascertained health factors are defined in S1 Table.

“High risk” was defined as the presence of current smoking (active or quit less than one month ago), or the presence of atherosclerotic cardiovascular disease (ASCVD) but reported non-adherence to an antiplatelet or a statin. Survey results for high risk patients were imported weekly into a high risk patient registry in REDCap (https://www.project-redcap.org). A nurse then reviewed these patients’ charts to categorize high risk behaviors and document interventions in the patient’s REDCap form.

The process for identifying high risk patients was reviewed with clinic managers and medical directors, who encouraged physicians to discuss smoking cessation with current smokers at their office visit. The nurse called current smokers and used motivational interviewing^18^ to encourage tobacco abstinence. Interest scales, confidence scales, and stages of change were determined from telephone interview. Both interest and confidence were scored on 0-10 Likert scales (0 = Not important at all / Not confident at all; 10 = Most important goal / 100% Confidence) in increments of 2. Stage of change was categorized as pre-contemplation, contemplation, preparation, action, or maintenance^19,20^. Patients who could not be reached were scored based on chart review of documented smoking cessation counseling while admitted at MGH. Patients who accepted support were then referred to various local and online tobacco cessation resources, and the phone intervention and referral outcome were documented in the patient’s chart.

For patients indicating they had ASCVD but were not on an antiplatelet or a statin, their charts were reviewed to confirm history of disease, medication prescription, and allergy or contraindication. High risk intervention category (prevalent ASCVD on Aspirin not statin, prevalent ASCVD on statin not Aspirin, or prevalent ASCVD not on Aspirin or statin) was recorded in the patient’s REDCap form. Patients with a documented reason for not being on the medication were categorized as Not Applicable, and then further categorized as: Intolerance documented, Taking a different antiplatelet, Statin contraindicated, ASA contraindicated, Started medication at office visit, or No history ASCVD. For patients without a prescription, the pharmacy and/or physician was contacted. Patients who were already prescribed the medication with no documented contraindications were called. If patients were taking a medication but unaware that this was a statin (or rarely, aspirin) they were marked as having a ‘Knowledge deficit.’ The patient’s cardiovascular disease specialist and primary care provider were notified of the findings via email. Eligible patients were offered referral to the Cardiovascular Prevention Center for a risk reduction visit.

The primary outcome was number of ideal factors for all patients. We compared the proportion suboptimal for each LS7 factor in patients with and without ASCVD using a two-sample proportion t-test. We also performed similar analyses comparing those with and without diabetes mellitus. In secondary analyses, we calculated a continuous LS7 score of 0-14, with each factor scored as 0, 1, or 2 for poor, intermediate, or ideal, respectively. We compared the continuous LS7 score between those with and without ASCVD using linear regression; we also pursued adjustment for age, sex, ethnicity, and clinic. Statistical significance was assigned at alpha = 0.05. In sensitivity analyses, among only participants who fully completed the survey, we evaluated the number of ideal factors in these patients, and compared LS7 score between those with and without ASCVD using linear regression.

The data was analyzed using the statistical software, R, version 3.4.2 (R Foundation for Statistical Computing, Vienna, Austria. URL http://www.R-project.org/).

## Results

5,950 patients under the care of cardiovascular or cerebrovascular specialists at MGH were surveyed in the outpatient setting using electronic tablets. Patients were distributed among MGH Cardiology (3,618, 60.8 %), Stroke (902, 15.2%), and Vascular Surgery 1,430 (24.0 %). Baseline characteristics of surveyed participants are provided in Table 1. The mean (standard deviation) age was 64 (15) years, 2,613 (44%) were female, and 4,836 (81%) were white. 58.5% (3,478) indicated the presence of ASCVD; of these 911 (26.2%) reported prior myocardial infarction. Survey completion rate was 42% (2,526) for LS7 questions.

**Table 1.** Baseline characteristics

| Characteristic | Value |
| --- | --- |
|  | N = 5,950 |
| Age (years) | 64 (±15) |
| Female | 2,613 (43.9%) |
| American Indian | 10 (0.2%) |
| Asian | 190 (3.2%) |
| Black | 151 (2.5%) |
| Hispanic | 202 (3.4%) |
| Pacific Islander | 3 (0.05%) |
| White | 4,836 (81.3%) |
| Recent smoker | 389 (6.5%) |
| Diabetes | 1,079 (18.1%) |
| ASCVD | 3,478 (58.4%) |
| Coronary artery disease | 1,598 (26.9%) |
| Peripheral arterial disease | 1,443 (24.2%) |
| Myocardial infarction | 911 (15.3%) |
| Stroke/TIA | 1,178 (19.8%) |
| Congestive heart failure | 322 (5.4%) |
| Atrial fibrillation | 808 (13.6%) |
| Congenital heart disease | 389 (6.5%) |
| Cardiology clinic | 3,618 (60.8%) |
| Stroke clinic | 902 (15.2%) |
| Vascular surgery | 1430 (24.0%) |
| LS7 question survey completion | 2,526 (42.4%) |

Categorical variables are presented as n (%) and continuous as mean (sd).
ASCVD = atherosclerotic cardiovascular disease; LS7 = Life's Simple 7; TIA = transient ischemic attack

Among all 5,950 surveyed patients, only 34 (0.6%) achieved ideal status for all 7 factors (Fig 1). 743 (12.6%) achieved ideal status for at least 5 of 7 factors. 2,600 (43.7%) achieved ideal status for 2 or fewer of 7 factors. 730 (12.3%) were ideal for all 3 health-related behaviors (i.e., diet, physical activity, and smoking). While 5,076 (85%) were not smoking, 4,350 (73%) and 2,907 (49%) were not ideal for diet and physical activity, respectively; in fact, 2,439 (41%) and 1,775 (30%) had poor status for diet and physical activity, respectively (Table 2). Additionally, 151 (2.5%) were ideal for all 4 quantitative cardiovascular risk factors. Among all LS7 components, diet had the highest prevalence of poor status responses 2439 (41%).

**Table 2:** Percent of patients ideal, intermediate, and poor for each LS7 factor

| LS7 Factor | Ideal | Intermediate | Poor |
| --- | --- | --- | --- |
| Diet | 1321 (22.2) | 1911 (32.1) | 2439 (41.0) |
| Physical activity | 2776 (46.7) | 1132 (19.0) | 1775 (29.8) |
| Smoking | 5076 (85.3) | 224 (3.8) | 389 (6.5) |
| LDL cholesterol | 1996 (33.5) | 976 (16.4) | 233 (3.9) |
| Blood pressure | 1554 (26.1) | 2550 (42.9) | 296 (5.0) |
| Glucose | 1940 (32.6) | 967 (16.3) | 679 (11.4) |
| BMI | 1999 (33.6) | 1836 (30.9) | 1567 (26.3) |

<sup>a</sup>BMI = body-mass index; LDL = low-density lipoprotein; LS7 = Life's Simple 7

**Fig 1.**
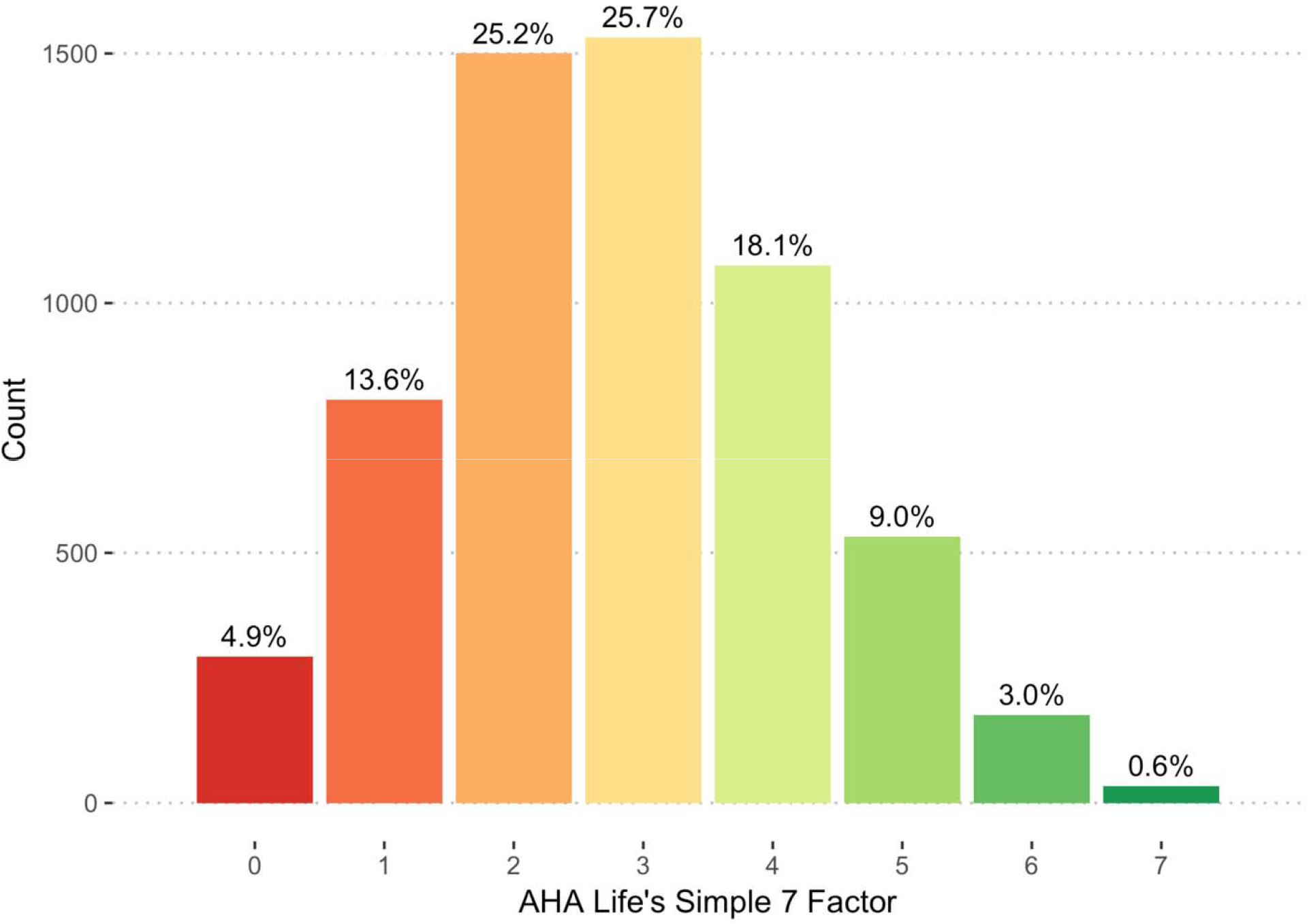
Distribution of ideal Life’s Simple 7 factors in patients cared for by cardiovascular or cerebrovascular specialists. Of the patients surveyed, ideal status was tabulated for each LS7 factor. A maximum score of 7 indicates ideal status across all factors. The distribution of this score across all surveyed patients is depicted. ^a^AHA = American Heart Association; LS7 = Life’s Simple 7

When weighted by poor (0), intermediate (1), or ideal (2) status across the 7 factors (maximum score 14), the mean (SD) score was 7.2 (2.9) (S2 Fig). Survey completion was a strong predictor of this score (Beta = 2.4, SE = 0.06, P value <0.001). Those who completed the survey fully tended to be mostly older (mean 66 versus 64 years; P value <0.001), white (84% versus 79%; P value <0.001), and with ASCVD (64% versus 54%; P value <0.001) (S2 Table). When only evaluating the 2,526 (42%) who fully completed the survey, the mean (SD) score was 8.9 (2.3) and only 30 (1.2%) were ideal across all 7 health-related behaviors (S3 Fig). In a multivariable model of demographics and prevalent cardiovascular diseases, diabetes mellitus and actively smoking were independently associated with a lower LS7 score; male sex, cardiology clinic, and stroke clinic were associated with a higher LS7 score (S3 Table).

Those with ASCVD had a similar proportion of individuals ideal for at least 5 of 7 health-related behaviors (444 (14.6%)) compared to those without ASCVD (298 (15.5%)) (P value = 0.49). Mean (standard deviation) LS7 score for patients with ASCVD was 7.5 (2.7) and 7.5 (2.6) for those without; when accounting for age, sex, ethnicity, and clinic, these scores were not significantly different (P value = 0.48) (S4 Fig). While a significantly higher proportion of individuals with ASCVD were ideal for LDL cholesterol, this was offset by higher proportion of suboptimal glucose, and suboptimal diet persisted at similar rates for those with and without ASCVD (S4 Table, Fig 2). Additionally, among those with ASCVD, 12% reported current smoking compared to 8% among those without ASCVD (P value <0.001). Among patients who fully completed the LS7 questions, we still did not observe a significant difference in LS7 score by ASCVD presence (S5 Fig). Similar results were found for those with diabetes mellitus compared to those without (S5 Table).

**Fig 2.**
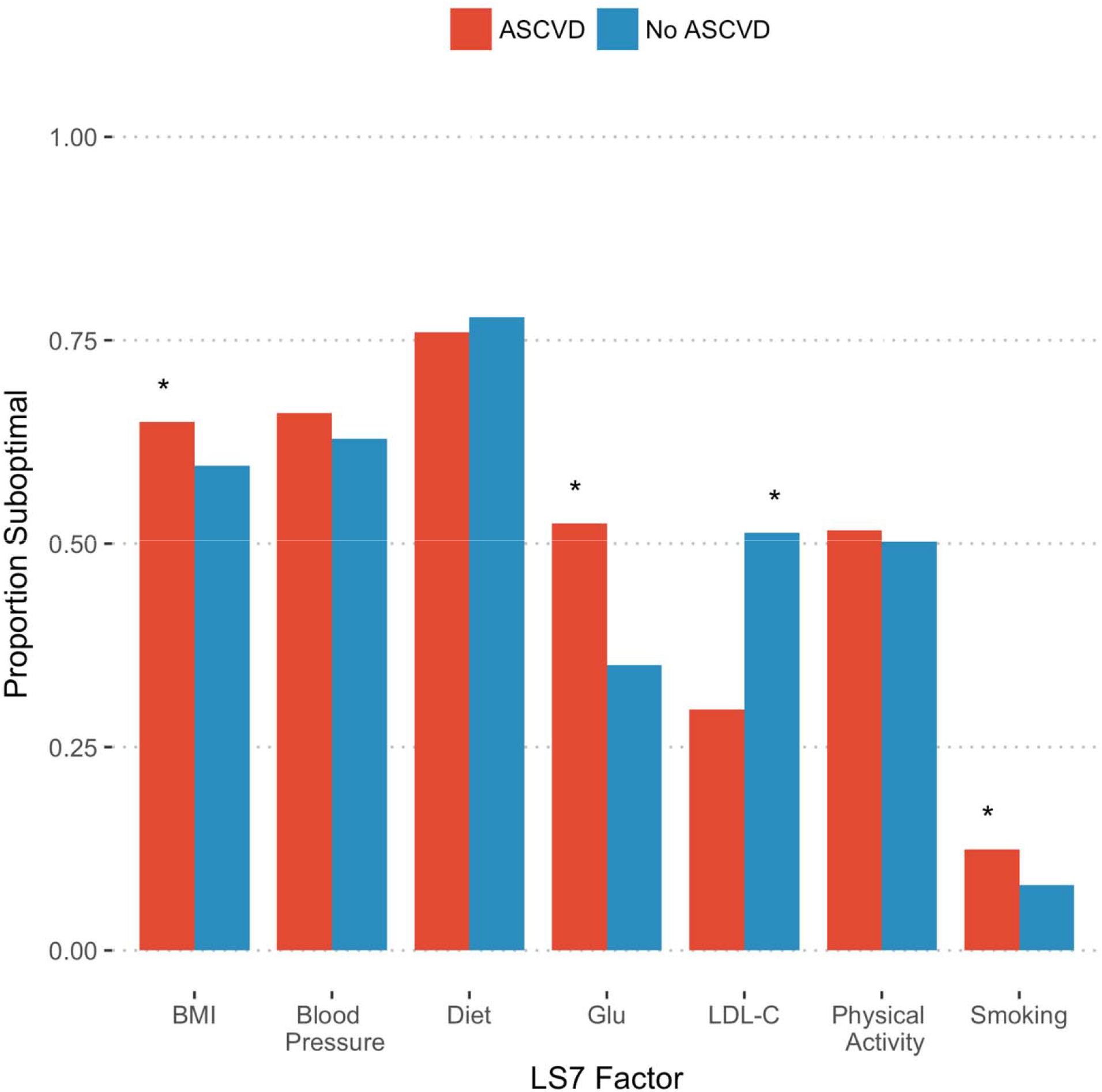
Proportion of patients with suboptimal LS7 factors by the presence of atherosclerotic cardiovascular disease. For each LS7 factor, the proportions of patients not ideal by ASCVD presence are depicted. Factors with at least nominally significant associations (P value < 0.05) are indicated with asterisks in the plot. ASCVD = atherosclerotic cardiovascular disease; BMI = body mass index; Glu = glucose; LDL-C = low-density lipoprotein cholesterol

Of those surveyed, 6% (389) indicated they were either active smokers or quit less than a month ago (Table 1). Fifteen additional smokers were identified when contacted about another risk factor. Of the smokers, the mean (standard deviation) age was 66 (15) years, 165 (40.8%) were female, and 273 (67.8%) had prevalent ASCVD. Out of 404 recent smokers, 94 (23%) were successfully counseled by phone, and 71 (18%) ultimately accepted additional support. Of those who accepted support, most accepted the nurse’s contact information (84%) for potential follow-up counseling and 38% accepted education materials regarding smoking cessation (S6 Table).

All 404 self-reported recent smokers’ charts were separately reviewed, and 289 were scored for three tobacco cessation readiness indices: interest, stage of change, and confidence. While 54% previously indicated in clinic notes high interest in smoking cessation (10 of 10 interest), most were in the pre-contemplative (35%) or contemplative (24%) stages of quitting. Nevertheless, 49% had high confidence (at least 8 of 10 confidence) in tobacco cessation (Table 3). Overall, the distributions were bi-modal with 26% displaying low interest (≤2 of 10) and 37% displaying low confidence (≤4 of 10). Of the smokers who were successfully contacted, mean confidence and interest scores were similar (P value = 0.27 and 0.06, respectively) when tobacco cessation counseling was provided versus clinician chart documentation (S7 Table).

**Table 3.**
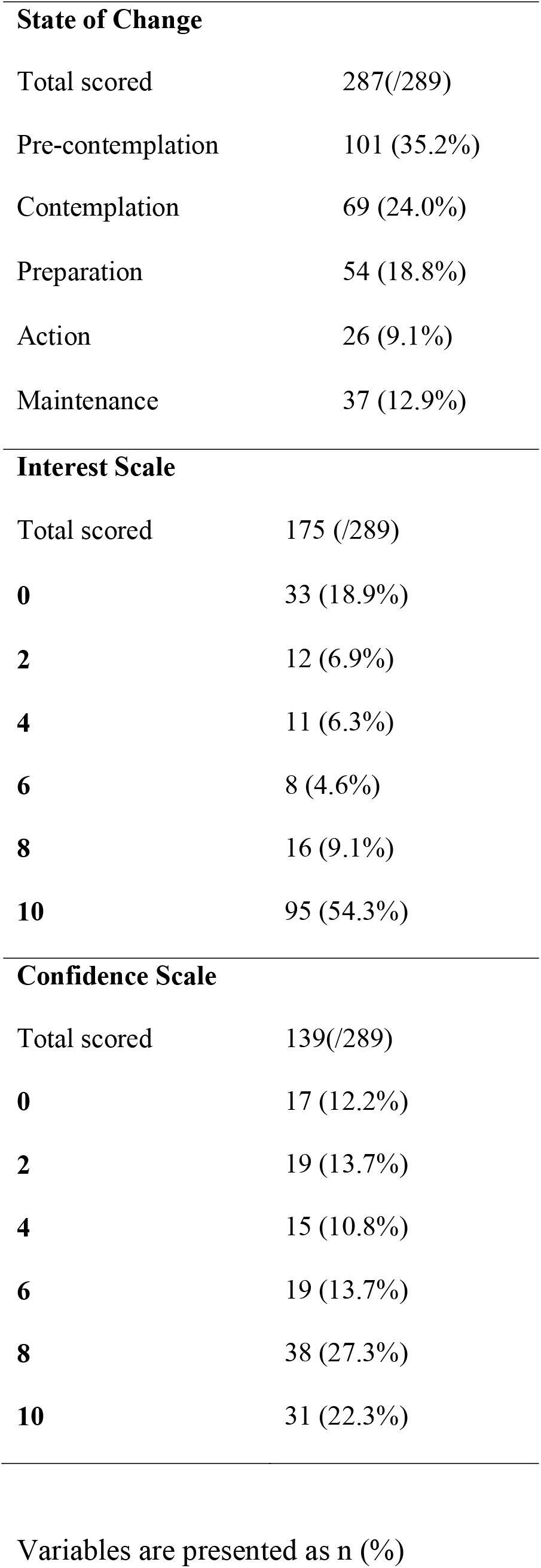
Tobacco cessation readiness indices of self-reported recent smokers

**State of Change**
|  |  |
| --- | --- |
| Total scored | 287(/289) |
| Pre-contemplation | 101 (35.2%) |
| Contemplation | 69 (24.0%) |
| Preparation | 54 (18.8%) |
| Action | 26 (9.1%) |
| Maintenance | 37 (12.9%) |
| **Interest Scale** |  |
| Total scored | 175 (/289) |
| **0** | 33 (18.9%) |
| **2** | 12 (6.9%) |
| **4** | 11 (6.3%) |
| **6** | 8 (4.6%) |
| **8** | 16 (9.1%) |
| **10** | 95 (54.3%) |
| **Confidence Scale** |  |
| Total scored | 139(/289) |
| **0** | 17 (12.2%) |
| **2** | 19 (13.7%) |
| **4** | 15 (10.8%) |
| **6** | 19 (13.7%) |
| **8** | 38 (27.3%) |
| **10** | 31 (22.3%) |

2 Variables are presented as n (%)

We identified patients with ASCVD who reported they were not taking a statin or an antiplatelet, and reviewed their charts to identify explanations for non-adherence. Of those with ASCVD, 868 (25%) reported not taking a statin or antiplatelet, or both. Based on chart review and contact with physician or pharmacy, non-adherence without contraindication was confirmed in 84 of 868 (10.1%), representing 2.4% patients with ASCVD (Table 4). Cardiovascular specialists and primary care doctors were contacted for these patients.

**Table 4.** Patients with ASCVD not taking a statin or antiplatelet or both

|  |  |
| --- | --- |
| Statin alone | 256 (29.5%) |
| Aspirin/clopidogrel alone | 381 (43.9%) |
| Both statin and aspirin/clopidogrel | 197 (22.7%) |
| Total | 834 |
| Physician, Pharmacy or Patient contacted | 439 (50.6%) |
| Intolerance documented | 148 (17.0%) |
| Aspirin contraindicated | 16 (1.8%) |
| Statin contraindicated | 10 (1.1%) |
| Taking a different antiplatelet | 145 (16.7%) |
| Started medication at office visit | 51 (5.9%) |
| Self-Reported Non-adherence | 84 (9.7%) |
| Knowledge deficit | 370 (42.6%) |
| No history of ASCVD | 290 (33.4%) |
| Successfully reconciled or contraindicated | 559 (64.4%) |

\*Total patients with ASCVD not taking a statin or aspirin or Plavix (or both) was 868; 834 were confirmed by the risk factor nurse

## Discussion

Through the assessment of patient-reported outcome measures of cardiovascular health factors and behaviors, we highlight gaps in cardiovascular disease preventive care for patients cared for by specialists at a tertiary academic medical center as well as strategies to begin to address these gaps. Only 1 in 100 patients seeking specialty cardiovascular care are optimal for the AHA’s LS7 and only 1 in 8 are optimal for health-related behaviors; even after ASCVD is manifest, with the exception of improved LDL cholesterol values, the prevalence of optimal LS7 factors remains largely similar. For key high-risk behaviors, recent smoking, non-adherence to guideline-directed medicines, and metabolic syndrome with poor diet and physical activity, we implemented scalable quality improvement interventions.

Our findings may have important implications for ASCVD risk mitigation at the individual- and population-level. First, despite receiving care from cardiovascular or cerebrovascular specialists, the prevalence of ideal status across all health-related factors and behaviors is similar to reports in the general population ^6-10^. Optimal status for these factors is associated with reduced cardiovascular disease risk in a dose-dependent manner^21^. As such, the American College of Cardiology and American Heart Association recommend the promotion of heart-healthy nutrition and physical activity behaviors for all adults to reduce cardiovascular disease risk^22^. Given the marked differences only in ideal LDL cholesterol values, emphasis in preventive cardiovascular medicine remains largely focused on pharmacotherapy-driven LDL cholesterol reduction. While this remains a highly effective strategy to reduce cardiovascular disease risk, sizeable persistent cardiovascular disease risks persist^23,24^. Our study highlights that persistent high risk health-related behaviors are highly prevalent, even when clinical ASCVD is recognized by patients and cardiovascular specialists.

Second, assessing patient-reported outcome measures not only provides a means for refining cardiovascular disease risk, but an opportunity to mitigate risk. Prior work has shown that cardiovascular risk communication to patients promotes the improvement of quantifiable risk factors^25,26^. Therefore, we provided a patient-facing report with an assessment of responses as well as advice tailored to survey responses. We also used patient-reported medication adherence to reconcile discrepancies with physicians and pharmacists. For example, approximately 1 in 10 patients with ASCVD reported not taking a statin or antiplatelet without contraindications in our study, which is similar to adherence rates reported in other clinical settings^27,28^. A recent study within the Veterans Affairs Health System closely linked statin non-adherence to future risk of death^29^. A recent systematic review concluded that the intensification of patient care interventions within healthcare systems can improve the short- and long-term adherence of cardiovascular disease risk-reducing medications^30^. Here, we leverage patient-reported non-adherence as an opportunity for additional patient, physician, and pharmacist engagement where necessary to promote adherence.

Third, scalable behavioral modification for cardiovascular disease prevention may be achievable through remote proactive strategies. Referral for telephone tobacco cessation counseling is associated with higher smoking cessation rates compared to standard take-home materials^31-33^. When patients call such services, efficacy is improved with proactive (i.e., additional outgoing calls after an index incoming call) versus reactive (i.e., only counseling from incoming calls)

counseling^34^. However, by leveraging systematic collection of patient-reported outcome measures, we are able to provide proactive counseling. Our data shows that we are able to contact a group of individuals unlikely to seek tobacco cessation counseling.

While our study has several strengths, there are important limitations. First, to maximally evaluate the value of patient-reported outcome measures, patients were asked to input medical history and quantitative metrics, including blood pressure, lipids, and glucose. To minimize errors, when laboratory values were recently measured within our health system, these values were provided to patients. Potential medical history misclassification does not influence the calculation or interpretation of the LS7 score. Second, while we did not observe differences in the LS7 score between those with and without ASCVD, those with ASCVD may have had poorer scores antecedent to ASCVD diagnosis. Unfortunately, even among those who participate in cardiac rehabilitation, a minority maintain an exercise regimen as long as six months later^35,36^. Longitudinal surveying is required to characterize the impact of ASCVD diagnosis on health-related behaviors; nevertheless, regardless of potential improvements, the vast majority remain suboptimal across health factors. Third, our single-arm study did not measure the clinical efficacy of assessing patient-reported outcome measures itself; randomized controlled trials are required to estimate risk reduction efficacy.

## Conclusions

A very small fraction of patients cared for by cardiovascular and cerebrovascular specialists at a large academic medical center are optimal for the AHA’s LS7. Systematic collection of patient-reported outcome measures to refine cardiovascular disease risk, communicate prevention strategies, and promote proactive cardiovascular disease prevention within a health system is feasible to address persistent large gaps in modifiable cardiovascular disease risk.

## Data Availability

Data is not available. Data was collected at Massachusetts General Hospital as a part of a quality improvement initiative and cannot be shared outside the institution.

## Acknowledgements

We wish to thank the patients and clinical providers who participated in this quality improvement initiative at the Massachusetts General Hospital.

